# Association between zinc deficiency and aorta stiffness in non-diabetic hemodialysis patients

**DOI:** 10.1101/2022.05.11.22274954

**Authors:** Kunihiro Ishioka, Sumi Hidaka, Naoki Fujiwara, Mizuki Yamano, Yasuhiro Mochida, Machiko Oka, Kyoko Maesato, Hidekazu Moriya, Takayasu Ohtake, Shuzo Kobayashi

## Abstract

**Objectives:** Zinc deficiency (Zn < 60 μg/dL) is known to play an important role for vascular calcification. However, little data is available regarding the association between zinc deficiency and aorta stiffness in dialysis patients. Thus, we studied the relationship between zinc deficiency and aorta stiffness in non-diabetic hemodialysis(HD) patients.

**Methods:** Of 150 patients receiving maintenance HD at our hospital, we included 79 non-diabetic HD patients (age: 70±11 years, 49 men) after excluding 71 diabetic HD patients. Zinc deficiency was defined as Zn <60 μg/dL during pre-HD blood sampling. The association between zinc deficiency and aorta stiffness was analyzed. Aorta stiffness was evaluated as brachial-ankle pulse wave velocity (baPWV). Other surrogate markers for cardiovascular complications were also measured.

**Results:** The zinc deficiency group (ZD group) included 45 patients (57.0%). Compared to the zinc non-deficiency group (ZND group), patients with ZD group were significantly older, higher levels of CRP and hypoalbuminemia. Moreover, they had significantly higher levels of baPWV, and lower levels of ankle-brachial pressure index (ABI) (p<0.05). After adjusting for hypoalubuminuria, and CRP, multivariate analysis showed that age and zinc level were independent predictors of baPWV.

**Conclusion:** The study suggested that zinc deficiency may be an independent risk factor for aorta stiffness, even after adjusting for malnutrition and inflammation.

## Introduction

Dialysis patients are characterized by a high risk of developing cardiovascular disease (CVD) events such as ischemic heart disease, cerebrovascular accident, and heart failure, and a low survival rate after CVD 1). Vascular calcification, which is the cause of CVD, progresses even before the introduction of hemodialysis. There are two types of calcification : atherosclerotic calcification lesions, which are mainly in the intima, and Mönkeberg-type calcification lesions, which are mainly in the tunica media. The latter is predominant in dialysis patients2), the tunica media calcification has been reported to be related to arterial stiffness.

The pulse wave velocity (PWV) of the aorta is a representative index of arterial wall stiffness and a predictor of the risk of CVD death and total mortality in dialysis patients3). Among them, brachial-ankle peak wave velocity (baPWV) is one of the most important surrogate markers for arterial wall stiffness, as it can be easily measured between the brachial joint and the ankle joint. High levels of baPWV have been known to be a prognostic factor of all-cause mortality and cardiovascular mortality. for hemodialysis patients4).

Zinc is one of the nine essential trace elements in humans, and is involved in the function of over 300 types of enzymes, cytokines, hormones, etc. It regulates the activation and signal transduction mechanisms involved in intracellular metabolism and cell response in nutrient metabolism and in the central nervous, immune, endocrine, digestive, and circulatory systems 5) 6).

It has benn reported so far that the zinc deficiency among patients undergoing hemodialysis (HD) was high at 40%–78% 7)8)9); the rates of zinc deficiency (Zn<60 μg/dL) and marginal zinc deficiency (60–80 μg/dL) in HD patients in Japan were 51.0% and 44.4%, respectively10). Thus, approximately 95% of HD patients in Japan satisfy the standard value of zinc deficiency10).

Recent reports indicate that zinc plays an important role in suppressing the calcification of vascular smooth muscle 11) 12). Vascular calcification is serious problem, leading to poor prognosis in terms of cardiovascular events, particularly in patients with chronic kidney disease13)14). However, to our knowledge, there are no reports on the association between zinc and aorta stiffness in dialysis patients.

Therefore, we investigated the association between serum zinc levels and aorta stiffness in HD patients excluding those with diabetes because of ever well-known strong risk factor for aorta stiffness.

## Materials and methods

### Subjects (Fig.1)

Of all 150 maintenance HD patients who were outpatients at the Shonan Kamakura General Hospital Dialysis Center, 71 diabetic patients were excluded, and 79 patients were included in the study (Fig.1). All patients gave informed consent, and local ethical committee approved the study (ethical No.TGE 01845-024) and was performed in accordance with the principles of the Declaration of Helsinki.

**Fig. 1.**
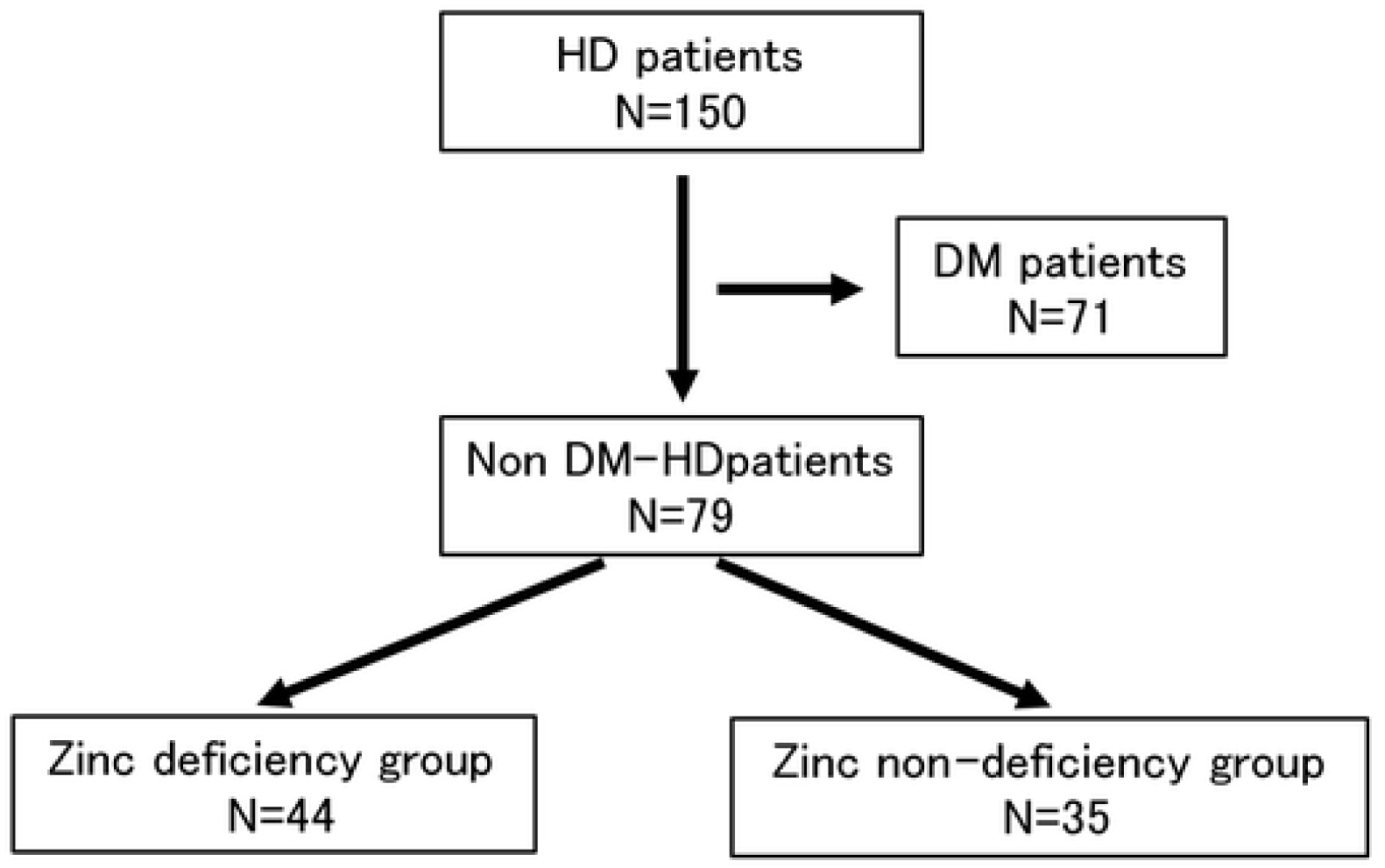
Classification between the zinc deficiency group and zinc non-deficiency groups HD, hemodialysis; DM, diabetes mellitus

### Zinc measurements and analysis items

Zinc measurements were conducted by promptly centrifuging the collected blood simultaneously with regular blood sampling (before dialysis at the beginning of the week, which was conducted after a 2-day interval), and the serum zinc levels were measured in our facility using the atomic absorption spectrophotometric method.

We analyzed relationships between serum zinc levels and the following: patient background (sex, age, body mass index, dialysis history, presence of diabetes), dialysis method (HD/online hemodiafiltration), regular blood sampling test items (39 in total), echocardiography parameters (ejection fraction, left ventricular mass index, E/e’, relative wall thickness, presence of valve calcification), and physiological tests (ankle-brachial index [ABI], toe-branchial index, baPWV, skin perfusion pressure[SPP]). The body mass index calculation used the estimated ideal weight(dry weight) at the time of regular blood sampling. Analyses were also conducted by dividing the group into those with Zn < 60 μg/dL, and those with Zn ≥ 60 μg/dL, which is one of the diagnostic criteria for zinc deficiency. baPWV/ABI was measured on non-dialysis days with Form PWV/ABI (OMRON Colin, Tokyo, Japan). SPP, which is used for detecting microcirculatory impairment, was measured at the same time with laser-doppler type device (PAD 4000®, Kaneka Co.,Ltd, Tokyo, Japan).

### Statistics

Continuous variables were compared using the Mann-Whitney U-test. The selection of explanatory variables for multiple regression analysis and binary logistic regression analysis were conducted using the stepwise variable increase/decrease method and variable increase method (likelihood ratio), respectively. The statistical analysis software used was JMP 10 (SAS Institute, JAPAN). The statistical significance level was set at p < 0.05.

## Results

### Patient background

The 79 (49 men, 62%) patients that were analyzed in this study had a median age of 70 (IQR 62–79) years, and a median hemodialysis duration of 6.9 (IQR 2.3–15.6) years.

#### Distribution of serum zinc

Fig 2 shows the frequency distribution of serum zinc levels in all patients. The median value was 58.4 (IQR 52.0–64.4; range 10.7–89.6) μg/dL. Forty five (57.0%) patients had zinc deficiency (<60 μg/dL), 32(40.5%)had latent zinc deficiency (60–80 μg/dL), and only 2 (2.5%) were within the normal range (80– 130 μg/dL) (Fig.2).

**Fig. 2.**
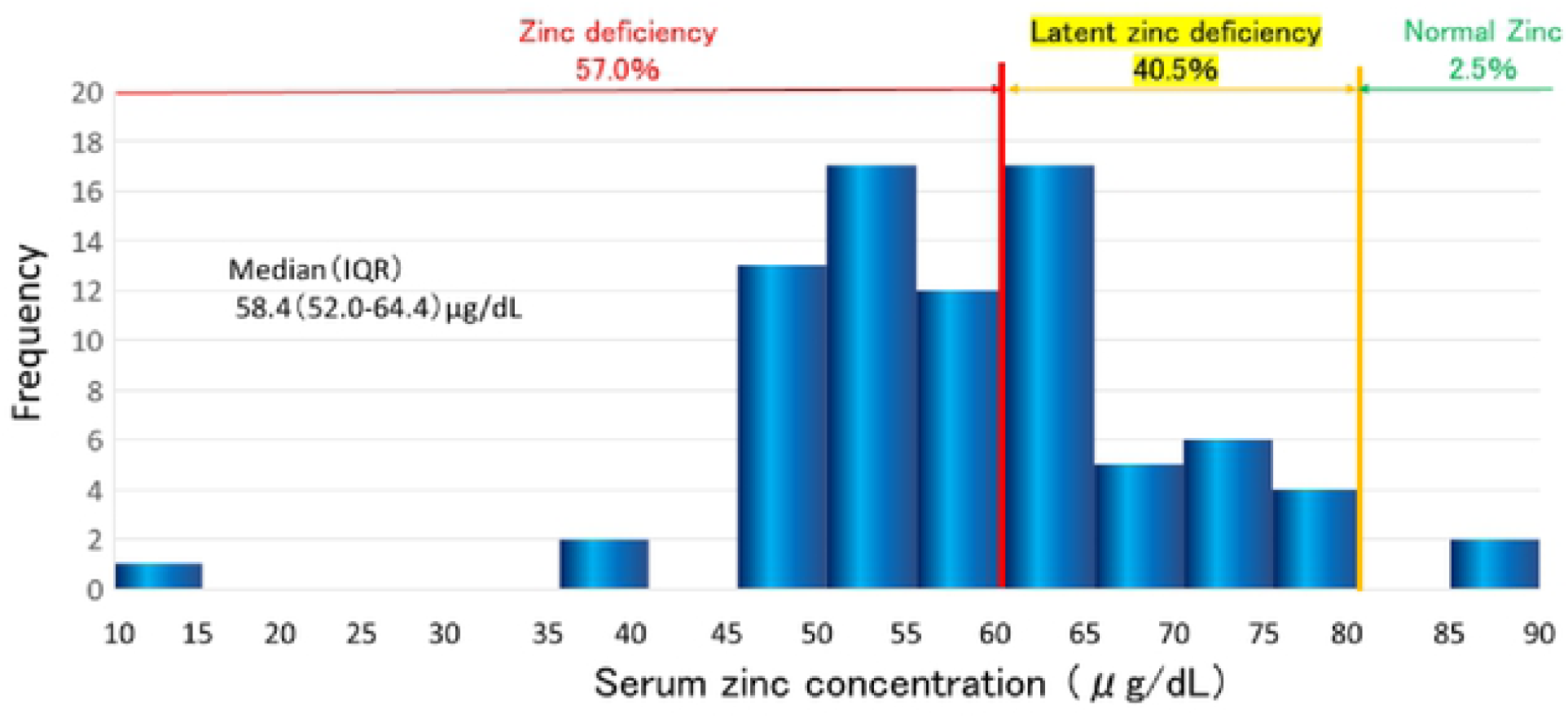
Distribution of serum zinc

#### Comparison between the zinc deficiency and non-deficiency groups

We divided the patients into the zinc deficiency group (ZD group) (< 60 μg/dL) and non-deficiency group(ZND group)(≥ 60 μg/dL) (zinc levels: 51.4±7.9 vs. 68.6±7.6 μg/dL, p<0.001) (Table 1 and 2). We compared the patients with hematology and biochemistry data and physiological examinations. The ZD group showed a significantly higher age (vs. ZND group: 73±11 vs. 67±12 years, p=0.033) and higher prevalence of ischemic heart disease (25.0% vs. 5.7%, p=0.016). Blood sampling tests showed that albumin and prealbumin levels were significantly lower in the ZD group (3.3±0.4 g/dL vs. 3.6±0.3 g/dL, p=0.0002; 24.3±8.8 mg/dL vs. 29.3±7.5 mg/dL, p=0.0030, respectively). Serum creatinine levels were also lower in the ZD group (9.1±2.5 vs. 10.5±2.8 mg/dL, p=0.022).

**Table 1.** Patient backgrounds of the zinc deficiency and non-deficiency groups

|  | Zinc deficiency (+) | Zinc deficiency (-) | P |
| --- | --- | --- | --- |
| N (%) | 44 (56) | 35 (44) |  |
| Sex (male) n (%) | 29 (66) | 20 (57) | 0.43 |
| Age (years) | 72(63-81) | 69(57-75) | 0.03 |
| Dialysis history (months) | 68(23-158) | 125(41-190) | 0.29 |
| Underlying disease |  |  |  |
| Chronic glomerulonephritis n (%) | 23 (52) | 15 (43) | 0.41 |
| Renal sclerosis n (%) | 5 (11) | 4 (11) | 0.99 |
| Polycystic kidney disease n (%) | 6 (13) | 6 (17) | 0.67 |
| Diabetes n (%) | 0 (0) | 0 (0) |  |
| Ischemic heart disease n (%) | 11 (25) | 2 (6) | 0.02 |
| Cerebrovascular disease n (%) | 4 (9) | 2 (6) | 0.57 |
| Hypertension n (%) | 34 (77) | 27 (77) | 0.99 |
| Dry weight (kg) | 54.3 (43.2-62.2) | 55.5 (48.0-67.1) | 0.14 |
| Systolic blood pressure(mmHg) | 144(120-153) | 144(129-154) | 0.52 |
| Diastolic blood pressure(mmHg) | 76(64-83) | 80(66-90) | 0.13 |

Values are presented as median (interquartile range; IQR) or as n (%)

**Table 2.** Blood sampling and Physiological test results of the zinc deficiency

|  | Zinc deficiency (+) | Zinc deficiency (–) | P |
| --- | --- | --- | --- |
| AST (U/L) | 16 (12-22) | 14 (11-18) | 0.24 |
| ALT (U/L) | 10 (7-17) | 10 (7-16) | 0.77 |
| TP (g/dL) | 6.2 (5.9-6.5) | 6.3 (6.1-6.6) | 0.31 |
| Albumin (g/dL) | 3.4 (3.1-3.6) | 3.6 (3.5-3.9) | <0.001 |
| Pre-albumin (mg/dL) | 23.5 (18.1-27.7) | 27.5 (23.1-32.1) | 0.003 |
| BUN (mg/dL) | 65.2 (54.1-71.7) | 62.9 (53.2-74.3) | 0.74 |
| Creatinine (mg/dL) | 9.33 (6.99-10.88) | 10.70 (8.38-13.07) | 0.02 |
| Na (mEq/L) | 140 (136-142) | 139 (138-142) | 0.23 |
| K (mEq/L) | 4.8 (4.3-5.4) | 4.9 (4.4-5.2) | 0.72 |
| Cl (mEq/L) | 107 (105-110) | 106 (104-109) | 0.88 |
| Ca (mg/dL) | 8.0 (7.5-8.7) | 8.4 (7.7-8.9) | 0.24 |
| P (mg/dL) | 5.6 (4.8-6.2) | 5.9 (4.8-6.8) | 0.24 |
| Ca*P (mg<sup>2</sup>/dL<sup>2</sup>) | 47.8 (36.3-53.9) | 49.4 (37.0-60.5) | 0.55 |
| Mg (mg/dL) | 2.3 (2.1-2.5) | 2.4 (2.0-2.5) | 0.57 |
| Fe (µg/dL) | 56 (36-80) | 69 (50-96) | 0.09 |
| Ferritin (ng/mL) | 95.5 (37.3-179.4) | 70.4 (24.7-177.6) | 0.61 |
| Zn (µg/dL) | 52.9 (48.9-55.9) | 64.6 (62.5-78.5) | <0.001 |
| Cu (µg/dL) | 95 (87-121) | 93 (80-107) | 0.07 |
| Transferrin (mg/dL) | 190 (162-204) | 191 (168-215) | 0.54 |
| CRP (mg/dL) | 0.260 (0.071-1.049) | 0.125 (0.040-0.335) | 0.05 |
| UA (mg/dL) | 7.0 (5.7-7.9) | 6.8 (5.9-7.6) | 0.95 |
| T. chol (mg/dL) | 166 (143-200) | 184 (155-213) | 0.21 |
| TG (mg/dL) | 91 (66-118) | 104 (88-128) | 0.11 |
| HDL-C (mg/dL) | 55.5 (41.7-67.7) | 50.2 (40.6-59.8) | 0.39 |
| LDL-C (mg/dL) | 86 (67-113) | 98 (70-122) | 0.22 |
| HbA1c (%) | 4.9 (4.6-5.3) | 4.9 (4.6-5.1) | 0.36 |
| Glycated albumin (%) | 15.8 (14.7-17.7) | 14.0 (13.0-15.5) | 0.002 |
| Pre-dialysis Glucose (g/dL) | 109 (95-133) | 100 (83-115) | 0.08 |
| β2MG (mg/L) | 30.1 (24.8-37.9) | 27.8 (22.6- 35.6) | 0.37 |
| intact PTH (pg/mL) | 149 (92-220) | 134 (88-155) | 0.71 |
| hANP (pg/mL) | 66.7 (32.5- 140.1) | 89.4 (49.3-160.5) | 0.13 |
| Hb (g/dL) | 10.7 (10.1-12.0) | 11.1 (10.7-11.9) | 0.21 |
| Ht (%) | 32.9 (31.4-37.5) | 34.1 (32.7-36.4) | 0.47 |
| Ret (%) | 1.3 (0.9-1.7) | 1.3 (1.1-1.9) | 0.30 |
| EF (%) | 60.6 (56.9-64.7) | 62.9 (58.7-66.1) | 0.11 |
| LVMI (g/m<sup>2</sup>) | 116.5 (95.1-139.3) | 112.3 (99.8-143.2) | 0.81 |
| E/e' | 12.0 (9.7-16.2) | 10.6 (7.0-15.1) | 0.21 |
| Valvular disease n (%) | 24 (55) | 23 (68) | 0.24 |
| Aortic valve calcification n (%) | 22 (50) | 21 (62) | 0.30 |
| Mitral valve calcification n (%) | 9 (21) | 11(32) | 0.23 |
| Aortic valve stenosis n (%) | 10 (23) | 4 (12) | 0.20 |
| Total ABI | 1.16 (1.10-1.25) | 1.25 (1.11-1.29) | 0.008 |
| baPWV(cm/s) | 1847 (1534-2241) | 1615 (1448-1903) | 0.01 |
| TBI | 0.71 (0.64-0.83) | 0.79 (0.67-0.87) | 0.06 |
| ABI<1.0 n (%) | 10 (23.8) | 4 (11.4) | 0.15 |
| ABI<0.9 n (%) | 7 (16.7) | 1 (2.9) | 0.04 |
| SPP (mmHg) | 77 (64-85) | 71 (63-89) | 0.68 |
| ABI<1.0 or SPP<50 n (%) | 14 (32.6) | 9 (26.4) | 0.56 |

Values are presented as median (interquartile range; IQR) or as n (%).
AST, aspartate transaminase; ALT, alanine transaminase; TP, total protein; BUN,
blood urea nitrogen; Ca, calcium; P, phosphate; CRP, C-reactive protein; UA, uric acid; T. chol, total cholesterol; TG, triglyceride; HDL-C, high-density lipoprotein-cholesterol; LDL-C, low-density lipoprotein-cholesterol; $\beta$ 2MG, $\beta$ 2 microglobulin; PTH, parathyroid hormone; hANP, human atrial natriuretic peptide; Hb, hemoglobin; Ht, hematocrit; Ret, reticulocytes EF, ejection fraction; LVMI, left ventricular mass index; ABI, ankle brachial index; baPWV, brachial-ankle pulse wave velocity; TBI, toe brachial index; SPP, skin perfusion pressure

The levels of baPWV were significantly higher in the ZD group (1958±748 vs. 1727±441, p=0.011) (Table 2). As shown in Fig 3, there is a significant correlation between baPWV and serum Zinc levels (R=0.298, P<0.05). ABI values are significantly lower in ZD group, compared with that in ZND group (1.12±0.19 vs. 1.21±0.15, p=0.008).

**Fig. 3.**
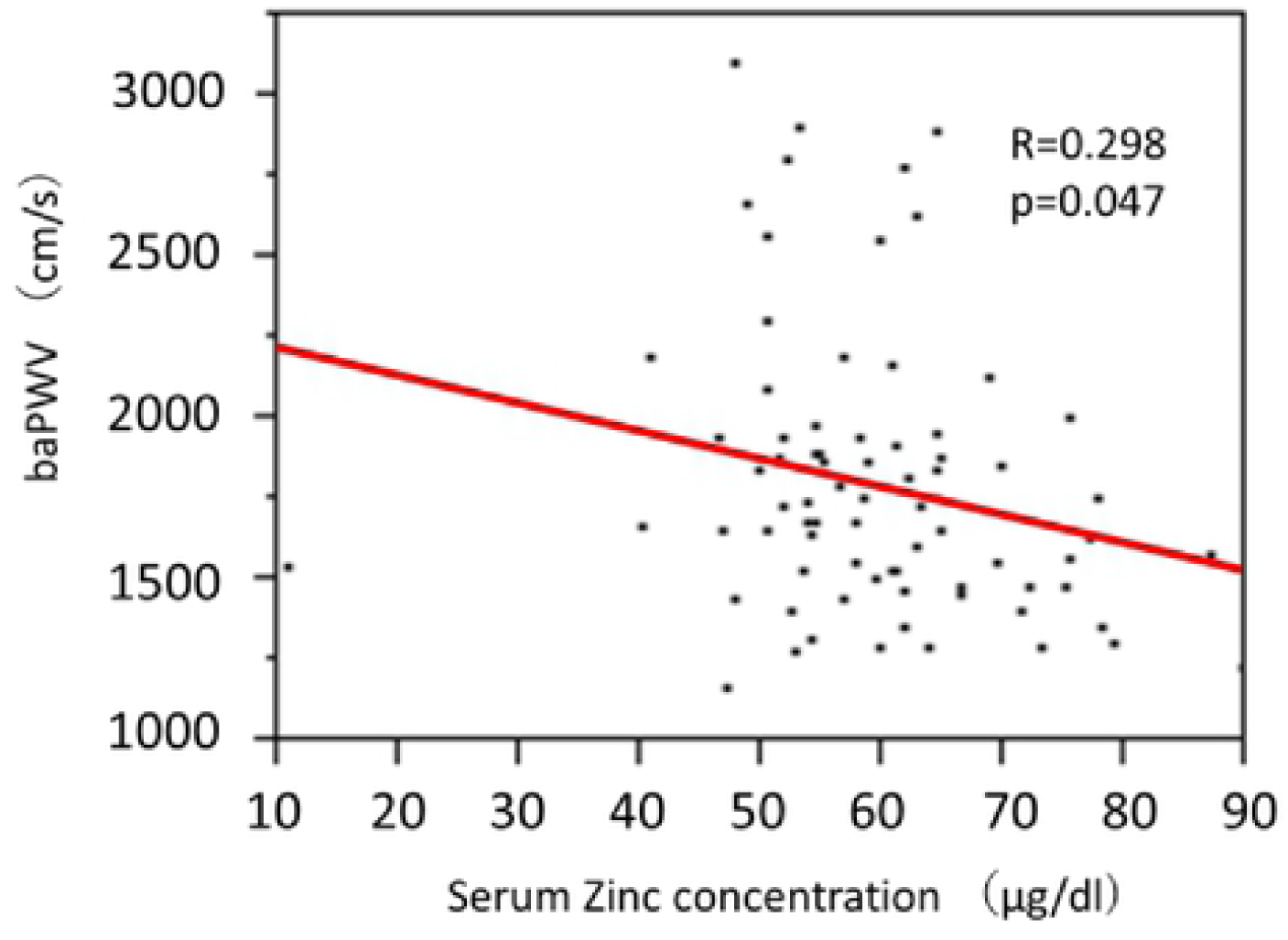
Relationship between serum zinc concentration and baPWV. serum zinc concentration and baPWV significantly and negatively correlated (r = 0.298, p = 0.047). baPWV, brachial-ankle pulse wave velocity.

#### Multivariate analysis

To examine factors associated with baPWV, a multivariate analyses was conducted with baPWV as the dependent variable and the results of blood test showing significant differences between the ZD group and ZND group as explanatory variables (Table 3). We found that zinc were independent predictors of baPWV (β= -9.85, t value= -2.00, p=0.047) as well as older age (Table 3).

**Table 3.**
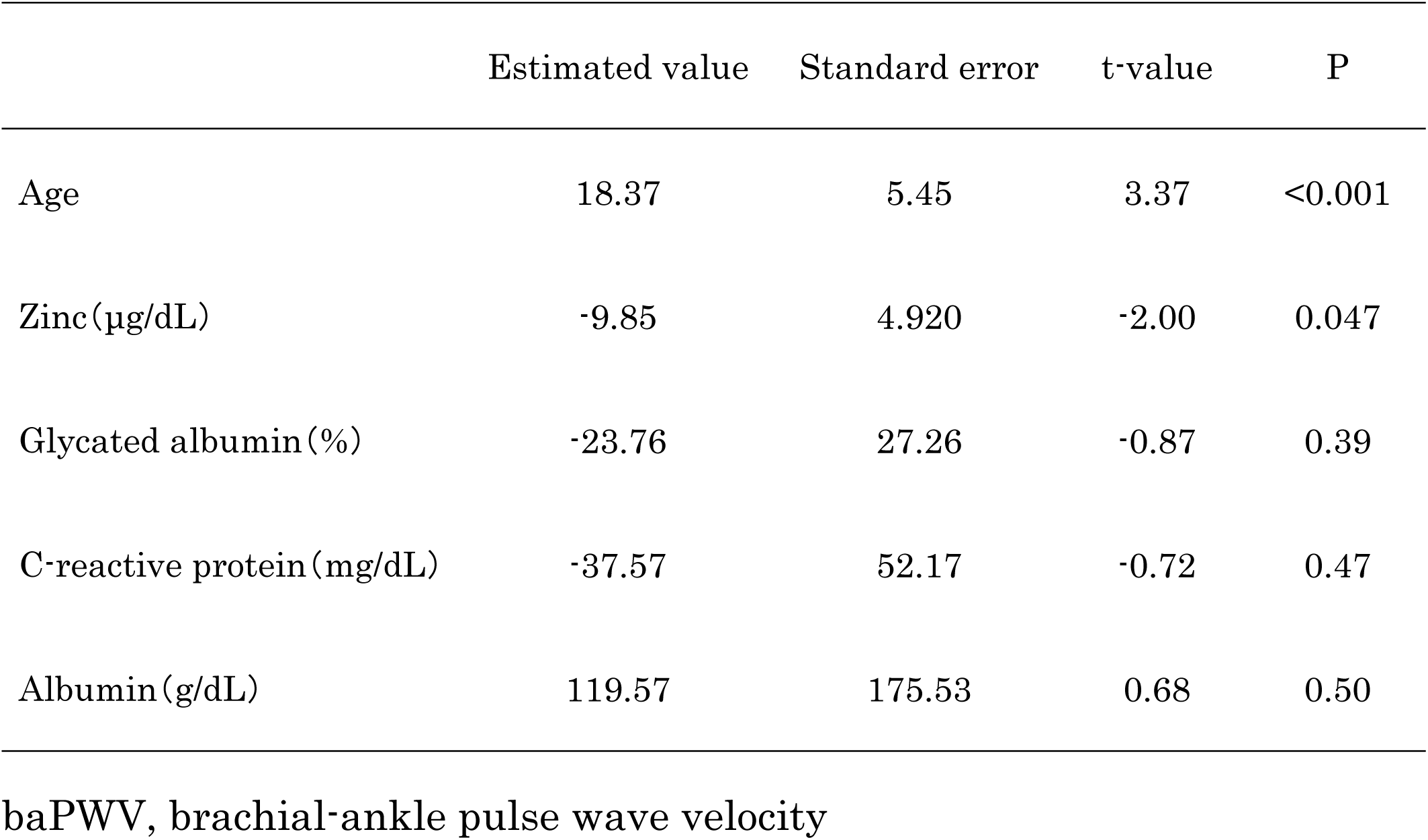
Multivariate analysis of predictors of baPWV

## Discussion

Our study clearly showed that zinc level was an independent predictive factor of baPWV, which is an indicator of aorta stiffness in HD patients. Risk factors for atherosclerosis and CVD in CKD patients include classic risk factors such as diabetes, old age, male sex, hypertension, and smoking, as well as the non-classic risk factors such as anemia, oxidative stress, inflammation, and malnutrition 15). Our present results indicate that hypozincemia is associated with aorta stiffness in HD patients.

The onset and progression of atherosclerosis and CVD begin from early stages of CKD. Atherosclerotic changes in the coronary arteries, particularly coronary artery calcification, are noted before the introduction of dialysis; when the estimated glomerular filtration rate (eGFR) drops below 45 mL/min/1.73 m2, both the prevalence and extent of calcification significantly increase14). Coronary artery stenosis has been frequently observed during the introduction of dialysis, affecting 53% of asymptomatic CKD patients and 83% of diabetic CKD patients 16).

Several studies have reported that zinc plays an important role in suppressing the calcification of vascular smooth muscle cells (VSMCs). Voelkl et al. reported that zinc sulfate 1) suppressed phosphate-induced calcification, 2) decreased the expression of mRNA (which is a bone formation marker that includes the expression of zinc-finger protein TNF-α-induced protein 3(TNFAIP3)), and 3) inhibited NF-kB activation and bone/cartilage reprogramming, and consequently suppress VSMC calcification by phosphate11). Therefore, serum zinc levels should be carefully maintained. Indeed, it is reported that increased dietary intake of zinc decrease 8% reduction of aorta calcification17).

In the present study, serum zinc deficiency was associated with aorta stiffness independent from serum albumin levels. The fact was similar to as the report that zinc deficiency was associated with 2-year mortality in HD patients18). Therefore, we need to consider the other factor for this association. Inflammation might be an important factor because malnutrition-inflammation-atherosclerosis (MIA syndrome) is well-known as a central axis19). In this regard, however, CRP values were not chosen as independent factor.

Peripheral arterial disease (PAD) is one of serious complications among HD patients 20). ABI is used as a screening test for the diagnosis of PAD. Also, vascular calcification plays an important role for the pathophysiology of PAD. The present study showed that ABI in ZD group was significantly lower than that of ZND group. Berard, et al show that in dialysis patients zinc deficiency had 3.4-fold increase in development of PAD21).

There are several limitations to this study. First, this was a cross-sectional observational study. Second, this study was conducted in a single facility and had a small sample size. Third, diabetic HD patients were excluded because of the greater risk factor of atherosclerosis, other risk factors for atherosclerosis in CKD patients were not sufficiently excluded. This study excluded diabetes, which is a major risk factor for aorta stiffness; however, classic risk factors such as hypertension and nutritional evaluation/lack of exercise were either not excluded or not incorporated into this study.

In conclusion, this single-center cross-sectional study showed that zinc levels in HD patients was an independent predictor of baPWV, which is an indicator of aorta stiffness. Further study is needed to clarify whether hypozincemia worsens the prognosis of HD patients.

## Data Availability

All relevant data are within the manuscript and its Supporting Information files.

## Acknowledgments

Not applicable.

## References

1) Foley RN, Parfrey PS, Sarnak MJ. Epidemiology of cardiovascular disease in chronic renal disease. J Am Soc Nephrol. 1998; 9(12 Suppl):S16–S23.

2) Gauthier-Bastien A, Ung RV, Larivière R, Mac-Way F, Lebel M, Agharazii M. Vascular remodeling and media calcification increases arterial stiffness in chronic kidney disease. Clin Exp Hypertens. 2014; 36(3): 173–180.

3) Shoji T, Emoto M, Shinohara K, Kakiya R, Tsujimoto Y, Kishimoto H, et al.: Diabetes mellitus, aortic stiffness, and cardiovascular mortality in end-stage renal disease. J Am Soc Nephrol. 2001; 12(10):2117–2124.

4) Kitahara T, Ono K, Tsuchida A, Kawai H, Shinohara M, Ishii Y, et al. Impact of brachial-ankle pulse wave velocity and ankle-brachial blood pressure index on mortality in hemodialysis patients. Am J Kidney Dis. 2005; 46(4): 688–696.

5) Prasad, AS. Zinc: Role in immunity, oxidative stress, and chronic inflammation. Curr Opin Clin Nutr Metab Care. 2009; 12(6): 646–652.

6) MacDonald, RS. The role of zinc in growth and cell proliferation. J Nutr. 2000; 130(5S Suppl): 1500S–1508S.

7) Dvornik S, Cuk M, Racki S, Zaputovic L. Serum zinc concentrations in the maintenance hemodialysis patients. Coll Antropol. 2006; 30(1): 125–129.

8) Lee SH, Huang JW, Hung KY, Leu LJ, Kan YT, Yang CS, et al. Trace Metals’ abnormalities in hemodialysis patients: Relationship with medications. Artif Organs. 2000; 24(11): 841–844.

9) Tonelli M, Wiebe N, Hemmelgarn B, Klarenbach S, Field C, Manns B, et al. Trace elements in hemodialysis patients: a systematic review and meta-analysis. BMC Med. 2009; 7: 25

10) Nagano N, Ito K, Oishi Y, Minami M,, Hayashi H, Tsunoda C, et al. Distribution of serum zinc levels in hemodialysis patients: Factors related to hypozincemia. J Jpn Soc Dial Ther. 2018; 51(6): 369–377, 2018 10.4009/jsdt.51.369

11) Voelkl J, Tuffaha R, Luong TTD, Zickler D, Masyout J, Feger M, et al. Zinc Inhibits Phosphate-Induced Vascular Calcification through TNFAIP3-Mediated Suppression of NF-kappaB. J. Am. Soc. Nephrol. 2018; 29(6): 1636–1648.

12) Chen Z, Gordillo-Martinez F, Jiang L, He P, Hong W, Wei X, et al. Zinc ameliorates human aortic valve calcification through GPR39 mediated ERK1/2 signalling pathway. Cardiovasc Res. 2021; 117(3): 820–835.

13) Kobayashi S. Cardiovascular events in chronic kidney disease (CKD)-an importance of vascular calcification and microcirculatory impairment. Renal Replacement Therapy. 2016; 2: 55.

14) Kobayashi S, Oka M, Maesato K, Ikee R, Mano T, Moriya H, et al. Coronary artery calcification, ADMA, and insulin resistance in CKD patients. Clin J Am Soc Nephrol. 2008; 3(5): 1289–1295.

15) Japanese Society for Dialysis Therapy: Clinical guidelines for the evaluation and the treatment of cardiovascular complications in hemodialysis patients. J Jpn Soc Dial Ther. 2011; 44(5): 337–425 10.4009/jsdt.44.337

16) Ohtake T, Kobayashi S, Moriya H, Negishi K, Okamoto K, Maesato K, et al. High prevalence of occult coronary artery stenosis in patients with chronic kidney disease at the initiation of renal replacement therapy: an angiographic examination. J Am Soc Nephrol. 2005; 16(4): 1141–1148.

17) Chen W, Eisenberg R, Mowrey WB, Wylie-Rosett J, Abramowitz MK, Bushinsky DA, et al. Association between dietary zinc intake and abdominal aortic calcification in US adults: Nephrol Dial Transplant. 2020; 35(7): 1171–1178.

18) Yang CY, Wu ML, Chou YY, Li SY, Deng JF, Yang WC, et al. Essential trace element status and clinical outcomes in long-term dialysis patients: a two-year prospective observational cohort study. Clin Nutr. 2012; 31(5): 630–6.

19) P Stenvinkel. The role of inflammation in the anaemia of end-stage renal disease. Nephrol Dial Transplant. 2001;16 Suppl 7: 36–40.

20) Ishioka K, Ohtake K, Moriya H, Mochida Y, Oka M, Maesato K, et al. High prevalence of peripheral arterial disease (PAD) in incident hemodialysis patients: screening by ankle-brachial index (ABI) and skin perfusion pressure (SPP) measurement. Renal Replacement Therapy; 2018: 4: 27.

21) Bérard AM, Bedel A, Le Trequesser R, Freyburger G, Nurden A, Colomer S, et al. Novel risk factors for premature peripheral arterial occlusive disease in non-diabetic patients: a case-control study: PLoS One. 2013; 8(3): e37882. doi: 10.1371/journal.pone.0037882.

